# Parental criticism and adolescent internalising symptoms: Using a Children-of-Twins design with power calculations to account for genetic influence

**DOI:** 10.1101/2020.05.07.20084319

**Authors:** Yasmin Ahmadzadeh, Thalia Eley, Laurie Hannigan, Cathy Creswell, Paul Lichtenstein, Erica Spotts, Jody Ganiban, Jenae Neiderhiser, Fruhling Rijsdijk, Tom McAdams

## Abstract

**Background:** Parental criticism is correlated with internalising symptoms in adolescent offspring. This correlation could in part reflect their genetic relatedness, if the same genes influence behaviours in both parents and offspring. We use a Children-of-Twins design to assess whether parent-reported criticism and offspring internalising symptoms remain associated after controlling for shared genes. To aid interpretation of our results and those of previous Children-of-Twins studies, we examine statistical power for the detection of genetic effects and explore the direction of possible causal effects between generations.

**Methods:** Data were drawn from two Swedish twin samples, comprising 876 adult twin pairs with adolescent offspring and 1030 adolescent twin pairs with parents. Parent reports of criticism towards their offspring were collected concurrently with parent and offspring reports of adolescent internalising symptoms. Children-of-Twins structural equation models were used to control for genetic influence on the intergenerational association between parental criticism and adolescent internalising.

**Results:** Parental criticism was associated with adolescent internalising symptoms after controlling for genetic influence. No significant role was found for shared genes influencing phenotypes in both generations, although power analyses suggested that some genetic effects may have gone undetected. Models could not distinguish directionality for non-genetic, causal effects between generations.

**Conclusions:** Parental criticism may be involved in psychosocial family processes in the context of adolescent internalising. Future studies should seek to identify these processes and provide clarity on the direction of potential causal effects.

---

Research suggests that parent-child relationships are important for adolescent adjustment (Laursen & Collins, 2009), during formative years of increasing autonomy and parent-child separation (De Goede, Branje, & Meeus, 2009). Parental criticism of adolescents has received attention as a possible mechanism relevant to their development of internalising symptoms (Asarnow, Tompson, Woo, & Cantwell, 2001; Frye & Garber, 2005; Nelemans, Hale Iii, Branje, Hawk, & Meeus, 2014; Silk et al., 2009). Parental criticism is characteristic of distressed or unsupportive interaction patterns within parent-child dyads (McCarty, Lau, Valeri, & Weisz, 2004), typically assessed using coded speech samples (e.g., Frye & Garber, 2005) or parent-report questionnaires (e.g., Nelemans et al., 2014). Associations between parental criticism and adolescent internalising may be driven by social interactions, such that parental criticism influences, or is influenced by, offspring symptoms (Hughes & Gullone, 2008). That is, parental criticism may contribute to offspring symptom development and/or reflect the parent’s reaction to child symptoms. However, this intergenerational association may also reflect influence by common causes, including genetics.

All human characteristics, including parenting behaviours and psychiatric symptoms, are influenced in part by genetics (Klahr & Burt, 2014; Plomin, DeFries, Knopik, & Neiderhiser, 2016; Polderman et al., 2015). Because offspring inherit genes from their parents, shared genes may at least partially account for correlations in their behaviour. Genes linked to complex behaviours typically have diffuse effects (Plomin et al., 2016), so the same genes could influence internalising symptoms during adolescence and parenting behaviours in adulthood. It is important that we understand the role of genes in families, to better understand the extent to which parent and offspring behaviours may be *causing* one another.

Genetic influence in families can be estimated and controlled for using causal inference designs, including the Children-of-Twins design (McAdams et al., 2014). Here, adult identical (monozygotic, MZ) and fraternal (dizygotic, DZ) twin pairs and their offspring are compared on any behaviours of interest. In families of MZ twins, offspring are equally related to both their parent and their aunt/uncle (genetic correlation=.50 for both; Figure1), although they share their immediate rearing environment only with their parent. In families of DZ twins, offspring are more genetically related to their parent (.50) than to their aunt/uncle (.25). So, if offspring behaviours are more correlated with aunt/uncle behaviours in MZ versus DZ families, genetic influence on the intergenerational correlation is inferred. If offspring behaviours are more correlated with parent versus aunt/uncle behaviours, then influence of the immediate rearing environment is inferred.

**Figure1.**
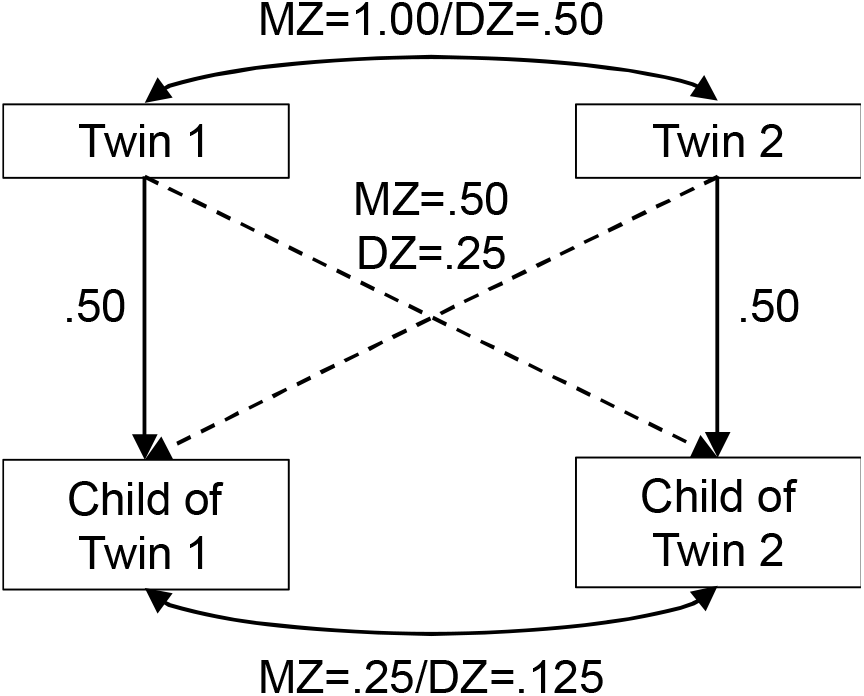
Genetic correlations between family members in the Children-of-Twins design for both monozygotic (MZ) and dizygotic (DZ) twin families

Existing Children-of-Twins research suggests that correlations between adolescent internalising symptoms and parents’ emotional overinvolvement (Narusyte et al., 2008), harsh punishment (Lynch et al., 2006) and parent-offspring relationship quality (Hannigan et al., 2018) are not at all attributable to genetic relatedness. The same is shown for adolescent somatic symptoms and parental criticism (Horwitz et al., 2015). These results are surprising, given the diffuse effects of genes linked to complex traits. Power calculations to accompany Children-of-Twins research would be useful, to clarify our ability to adequately detect and control for genetic influence in each analysis. These have not been presented before, so existing findings remain difficult to critically interpret. Further, only one of the aforementioned papers included analyses to explore the possible direction of causation (for non-genetic associations) between generations (Narusyte et al., 2008). Results suggested a stronger influence of child internalising problems on parents’ emotional overinvolvement, compared to the reverse, although results were undermined by uncertainty regarding methodology and statistical power.

Statistical power in Children-of-Twins designs depends on several factors, each of which can limit ability to detect genetic influence in parent-child associations. Factors include the magnitude of the parent-child correlation; proportion of the correlation attributable to genetic relatedness; magnitude of relatedness in avuncular pairs (Figure1 dashed lines); sample size; ratio of identical and fraternal twins; heritability of the parent and offspring behaviours; and power to detect heritability within each generation (McAdams et al., 2018; Verhulst, 2017). These factors vary across samples and variables. When power is not sufficient to detect genetic influence in Children-of-Twins analyses, intergenerational covariance appears attributable to non-genetic factors (McAdams et al., 2018). Previously published Children-of-Twins studies may have been underpowered, resulting in the overestimation of non-genetic effects. This has not been addressed in previous research.

We used a Children-of-Twins design to examine whether the association between parental criticism and adolescent offspring internalising symptoms remained significant after we account for possible genetic influence. We explore statistical power in our sample for the detection of genetic influence. Where evidence was found for non-genetic effects between parents and offspring, we sought to identify a causal direction between generations.

## Methods

### Participants

Data from two samples were combined for an extended Children-of-Twins design (as done elsewhere e.g., Hannigan et al., 2018; Narusyte et al., 2008): (1) a Children-of-Twins sample where the twins were adult parents with adolescent offspring, and (2) a sample of adolescent twin pairs with one parent per pair.

### Children-of-Twins sample

Data were drawn from the Twin and Offspring Study in Sweden (TOSS; Neiderhiser & Lichtenstein, 2008), comprising 387 MZ and 489 DZ twin families. Families consisted of same-sex adult twin pairs, with one genetically related adolescent child per twin. Each twin had been cohabiting with a partner (usually spouse) for a minimum of five years. Cousins (the offspring) were same sex and did not differ in age by >4 years. 37% of twins and 52% of offspring were male. Mean ages were 44.8 years for twins (SD=4.9; range 32-60) and 15.7 years for offspring (SD=2.4; range 11-22). In sensitivity analyses we excluded data from families where either offspring was >19 years (n=25), because these offspring could be considered young adults and less exposed to parental criticism.

### Adolescent twin sample

Data were drawn from Wave3 of the Swedish Twin Study of Child and Adolescent Development (TCHAD; Lichtenstein, Tuvblad, Larsson, & Carlstrom, 2007). Wave3 was chosen to match the adolescent ages and measures in the TOSS. The sample comprised 416 MZ and 614 DZ (299 same-sex DZ) adolescent twin pairs. One parent was included per twin pair. 49% of twins and 13% of parents were male. Twins had a mean age of 16.7 years (SD=0.47; range 15-17).

## Ethical Considerations

Written consent was obtained from all participants and ethical approval from the Institutional Review Boards of the home institutions concerned (Lichtenstein et al., 2007; Neiderhiser & Lichtenstein, 2008).

## Measures

### Parental Criticism

Parents reported on critical perceptions and behaviours towards their adolescent offspring, using the validated 10-item critical remarks subscale of a validated Expressed Emotion measure (Hansson & Jarbin, 1997). Self-reported questionnaires of parental criticism are useful in research to assess parents’ own view of their feelings and experiences with the child. Data collection and coding costs less compared to interview assessments (Hale et al., 2011). Parents responded using a five-point Likert scale to indicate how often they agreed with each critical statement, e.g., “S/he makes me irritated”, “It’s hard for us to get along” and “I find faults with him/her”. Cronbach’s alpha was >.85 in both samples (TableS1).

### Adolescent Internalising Symptoms

Adolescent internalising symptoms were reported by parents and adolescents using the 30-item Internalising scale of the Child Behaviour Checklist and Youth Self-Report, respectively (Achenbach, 1991a, 1991b). These corresponding parent and child assessments use the same three-point Likert scale and were moderately correlated in both samples (*r=*.36/.42 in TOSS/TCHAD). The internalising scale sums syndrome scales for anxious/depressed, somatic and withdrawn behaviours, which share most of their genetic aetiology during adolescence (Waszczuk, Zavos, Gregory, & Eley, 2014). Cronbach’s alpha was ≥.80 in both samples (TableS1). Parent and adolescent reports capture adolescent internalising phenotypes from different perspectives, with different biases associated with each method (De Los Reyes & Kazdin, 2005). We derived composite mean scores, to incorporate all available information. Including parent reports for all study variables could inflate the parent-offspring correlation via shared method variance. Therefore, we also conducted supplementary analyses using only self-reports for adolescent internalising.

## Analysis

The effects of child age and sex were regressed out from all variables. Variables were then log transformed to correct for skew (see TableS1) and standardised. Structural equation models were fitted using maximum likelihood estimation in the R (version 3.6.1) programme OpenMx (version 2.15.5; Neale et al., 2016). Confidence intervals were calculated using profile likelihood-based intervals, as provided by OpenMx. Data from different family types (comprising MZ or DZ, adult or adolescent twins) were specified separately in a single model (Figure2 shows the partial path diagram; FigureS1 shows full model specification). This model derived estimates for three sets of influences: (1) genetic and environmental influences on parental criticism, (2) covariance between the generations that is attributable to genetic versus non-genetic influences, and (3) genetic and environmental influences on offspring internalising symptoms.

**Figure2.**
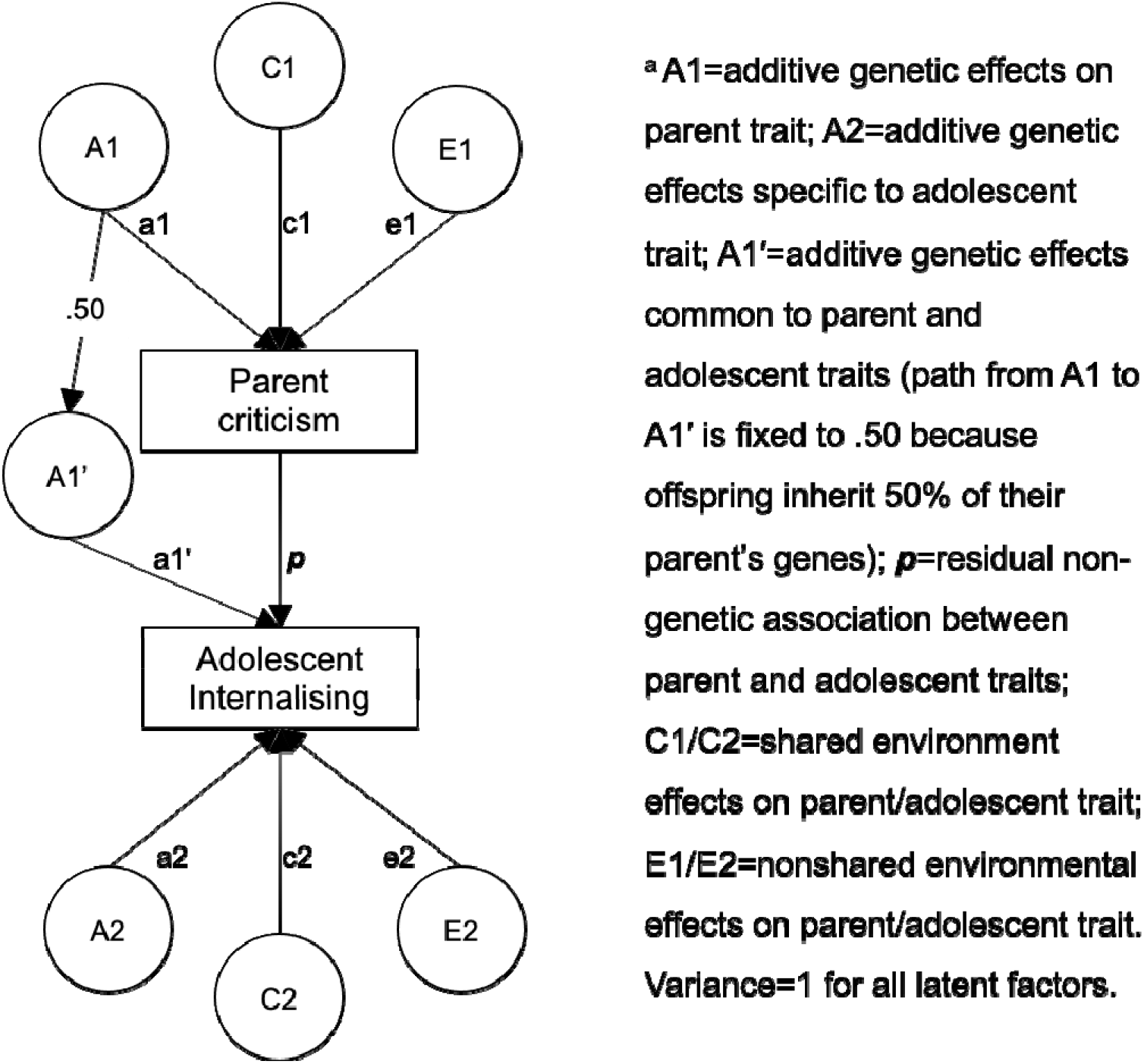
Partial path diagram for the Children-of-Twins structural equation model

Genetic and environmental influences on parental criticism were estimated using adult twin pairs, based on knowledge that genetic sharing is higher between MZ versus DZ twins (Figure1). As in the regular twin design (Polderman et al., 2015), we quantified genetic (A1), common environmental (C1) and non-shared environmental (E1) influences.

Intergenerational *covariance* was decomposed using comparisons of avuncular and parent-offspring correlations for MZ and DZ twins with children. Genetic sharing is higher for avuncular pairs in MZ versus DZ families (Figure1). This difference allowed us to estimate the relative contribution of genetic influence on the intergenerational association. Genetic covariance was modelled via A1’ (Figure 2). Any residual intergenerational association was estimated via *p*, capturing non-genetic influence. Non-genetic influence represents all pathways outside of parent-to-child genetic transmission. This includes environmentally mediated influence of heritable parent behaviours on child behaviour and/or of heritable child behaviours on parent behaviour (evocative gene-environment correlation).

Comparing correlations between the offspring of MZ and DZ twins (cousins) allowed us to estimate genetic (A2) and non-shared environmental (E2) influences on offspring internalising symptoms. This is because cousins have higher genetic sharing in MZ versus DZ families (Figure1). However, relatively low genetic sharing between cousins means that genetic influences on offspring traits can go undetected, due to low power. Because cousins do not share a home environment, they cannot inform on the influence of common environments on offspring traits (C2). Therefore, the adolescent twin dataset was used to additionally inform on the aetiology of offspring internalising symptoms. Twin pairs have higher genetic sharing compared to cousins, so yield greater power to detect A2, and can inform estimates of C2.

Parental criticism in the adolescent twin dataset contributed to estimates of the phenotypic parent-child correlation, boosting the number of parent-child correlations going into the model. These data did not inform on the decomposition of parent trait aetiology or intergenerational covariance. The same parent reported on their criticism of each twin; therefore, A1 and C1 correlations were fixed to unity for parents of adolescent twins (FigureS1b-c). Our approach was similar to that taken in previous studies combining adult and child twin datasets (e.g., Narusyte et al., 2008), with novel distinctions detailed in the Supporting Information.

The significance of model parameters in our complete model was tested by creating sub-models where paths were consecutively fixed to zero. χ^2^ difference tests and Akaike’s Information Criterion (AIC) were used to assess whether sub-models yielded significantly worse fits to the data. All models were re-run using only adolescent self-reports for child internalising symptoms.

### Power to detect genetic influence

Accurate estimation of A1’ (genetic contribution to the parent-child correlation) relies upon adequate statistical power. Estimation of the non-genetic *p* pathway is inflated in models with inadequate power to detect A1’, because *p* is the residual parent-offspring covariance after accounting for A1’. Therefore, we examined statistical power in our sample to detect an A1’ pathway of varied magnitudes. We simulated data to match our sample characteristics and systematically altered the strength of the A1’ pathway, noting the corresponding change in observed statistical power to detect it. We chose to conduct power analyses post-hoc, because it is difficult to estimate power to detect a single parameter a priori in large models when all parameters can affect one another (we estimate 8 paths concurrently) in unpredictable ways (Verhulst, 2017).

### Exploring the direction of causation

In Children-of-Twins models, the direction of non-genetic effects is usually modelled as running from parent-to-child, although any child-to-parent effects are also captured. To distinguish a causal direction for non-genetic effects, we ran unidirectional and bidirectional Children-of-Twins causal models. We followed protocol introduced by Narusyte et al. (2008; including our corrections to model specification) and Heath et al. (1993). Further details are provided in the supplementary materials.

## Results

TableS1 shows descriptive statistics for all measures. Table1 shows that correlations in parental criticism for adult twins were greater for MZ (.26) compared to DZ (.14) pairs, suggesting influence of parents’ genes on parents’ criticism. Parent-child correlations (.29, set to be equal across family types) were over twice the magnitude of avuncular correlations in MZ (.12) and DZ (.06) families. This shows stronger correlations between individuals who lived together, indicating influence of their immediate family environment. The avuncular correlations in MZ versus DZ twin families were not significantly different (Table1). For adolescent pairs, correlations in internalising symptoms were greater for MZ (.60) versus DZ (.31) twins; and cousins in MZ (.17) versus DZ (.09) families. This suggests influence of adolescents’ genes on adolescents’ internalising symptoms. Results were unchanged following exclusion of data from 25 families who had offspring aged 19–22 years. A similar pattern of results was found in models using only self-report data for adolescent internalising, although intergenerational correlations were smaller (TableS2).

**Table1.**
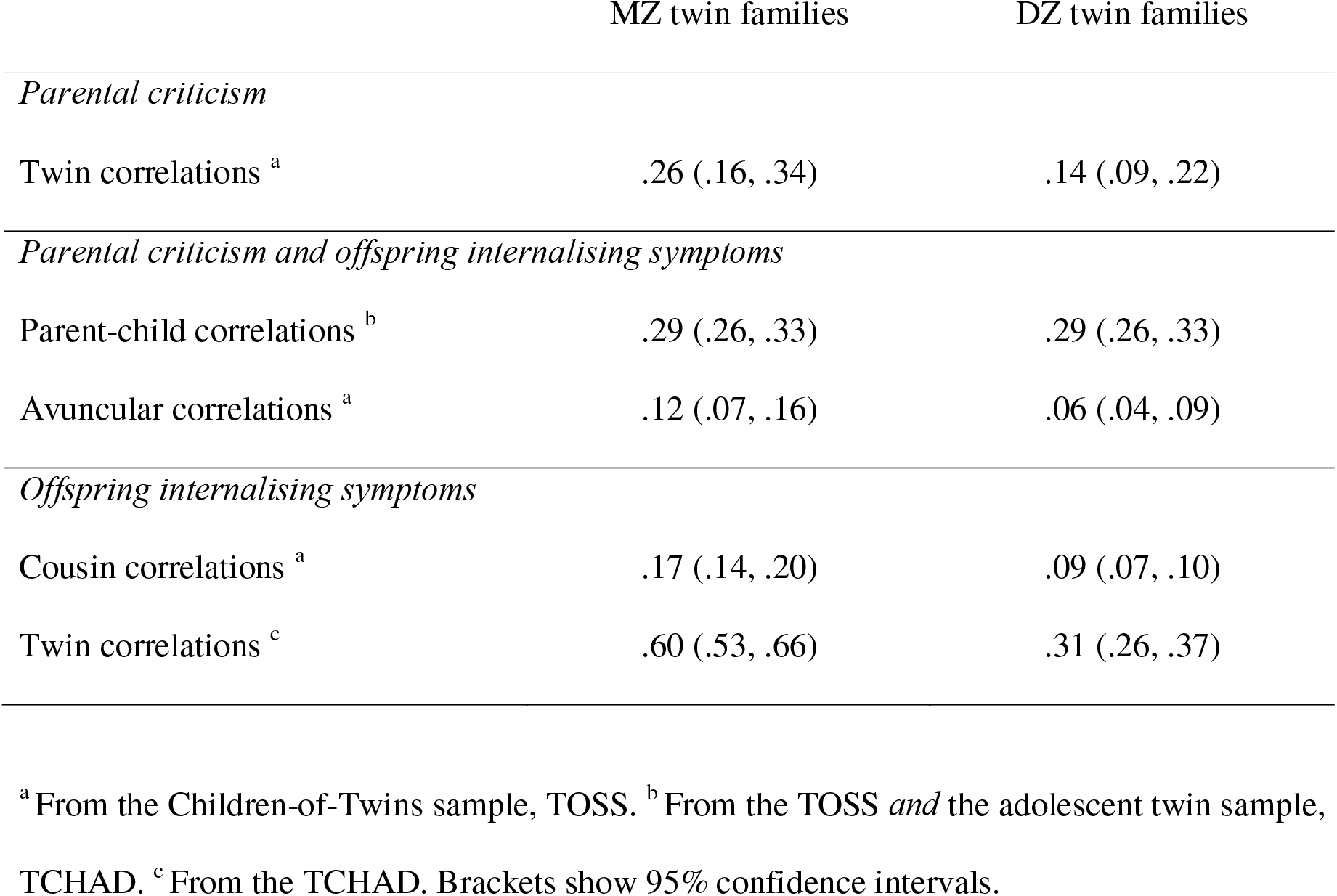
Correlations in monozygotic (MZ) and dizygotic (DZ) families

### Model Fitting

Model results are depicted in Figure3. C1 and C2 did not account for measure variance.^1^ Their exclusion resulted in a more parsimonious model, without significant changes to model fit (TableS3). Heritability of parental criticism and adolescent internalising symptoms were estimated at 26% and 53% respectively. For the intergenerational association, A1’ was not significantly different from zero (β=√.05, 95% CI .00-.41). The residual, non-genetic pathway (*p*; presented as a standardised beta estimate for direct comparison with the phenotypic parent-offspring correlation) was positive and significant (β=.24, 95% CI .18-.30). Subsequent nested models reinforced this observation. Specifically, the intergenerational association could be adequately explained by a model excluding genetic influence, but not by a model including only genetic influence (TableS3). Consistent conclusions were drawn from analyses using self-report data for adolescent internalising (FigureS2).

**Figure3.**
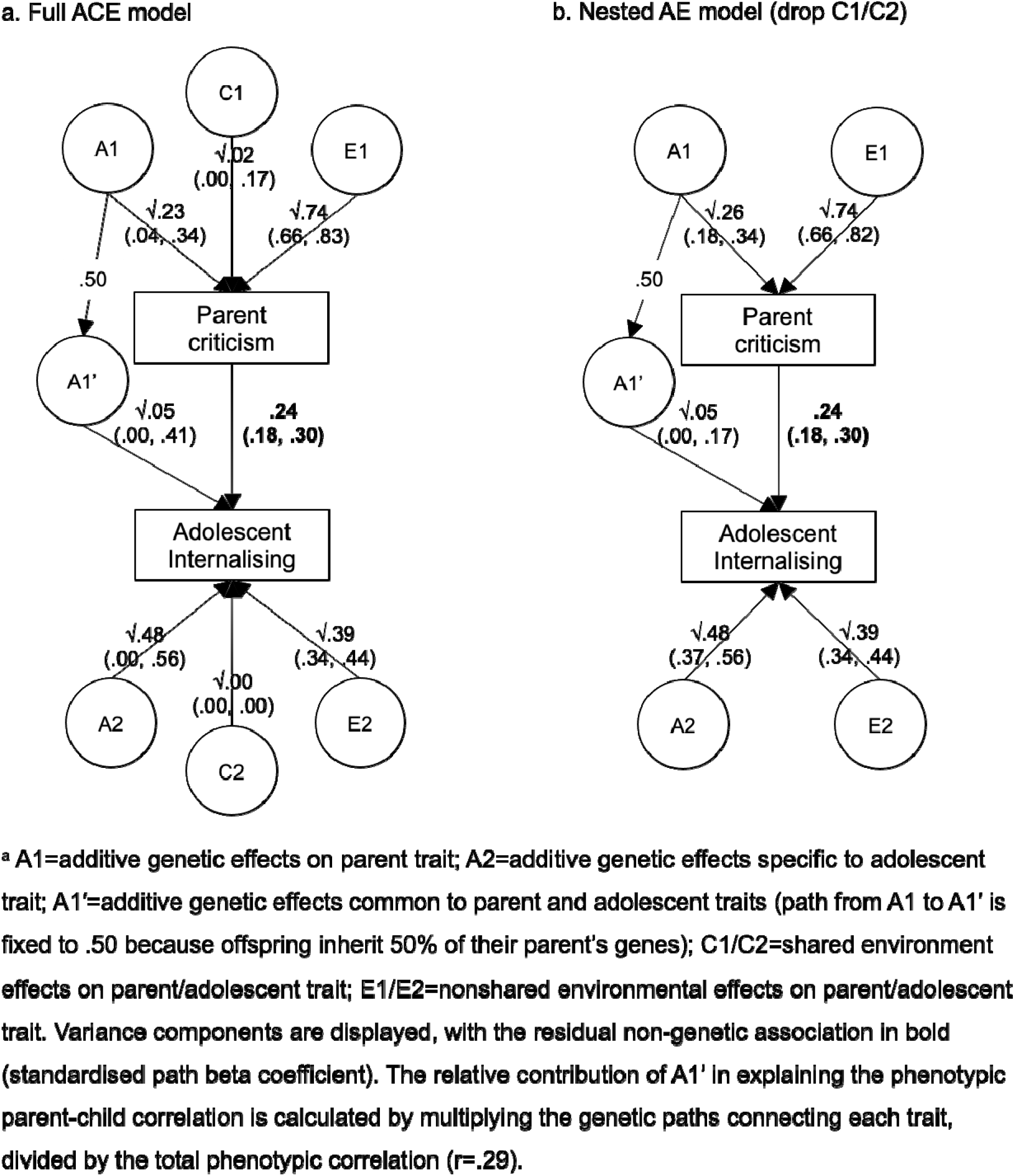
Model results, showing the association between parental criticism and offspring internalising symptoms ^a^

### Power to detect genetic influence

To evaluate statistical power in our sample to adequately detect genetic influence, we simulated a dataset to match the parameters in our final model. We examined how great the variance explained by A1’ must be to harness ≥80% power to detect it, keeping total parent and child heritability estimates and parent-child covariance static (Table2). There was ≥80% power to detect an A1’ estimate of .09 or greater (Table2, bold row). This would account for 26% of the phenotypic parent-child correlation. Therefore, results suggest that *at least* 74% of the association between parental criticism and offspring internalising symptoms was not attributable to genetic influence in this sample. The small A1’ estimate in Figure3b *could* be a true finding for genetic influence on parent-offspring covariance (accounting for 19% of the phenotypic association),^2^ but statistical power was not sufficient to confirm it.

**Table2.**
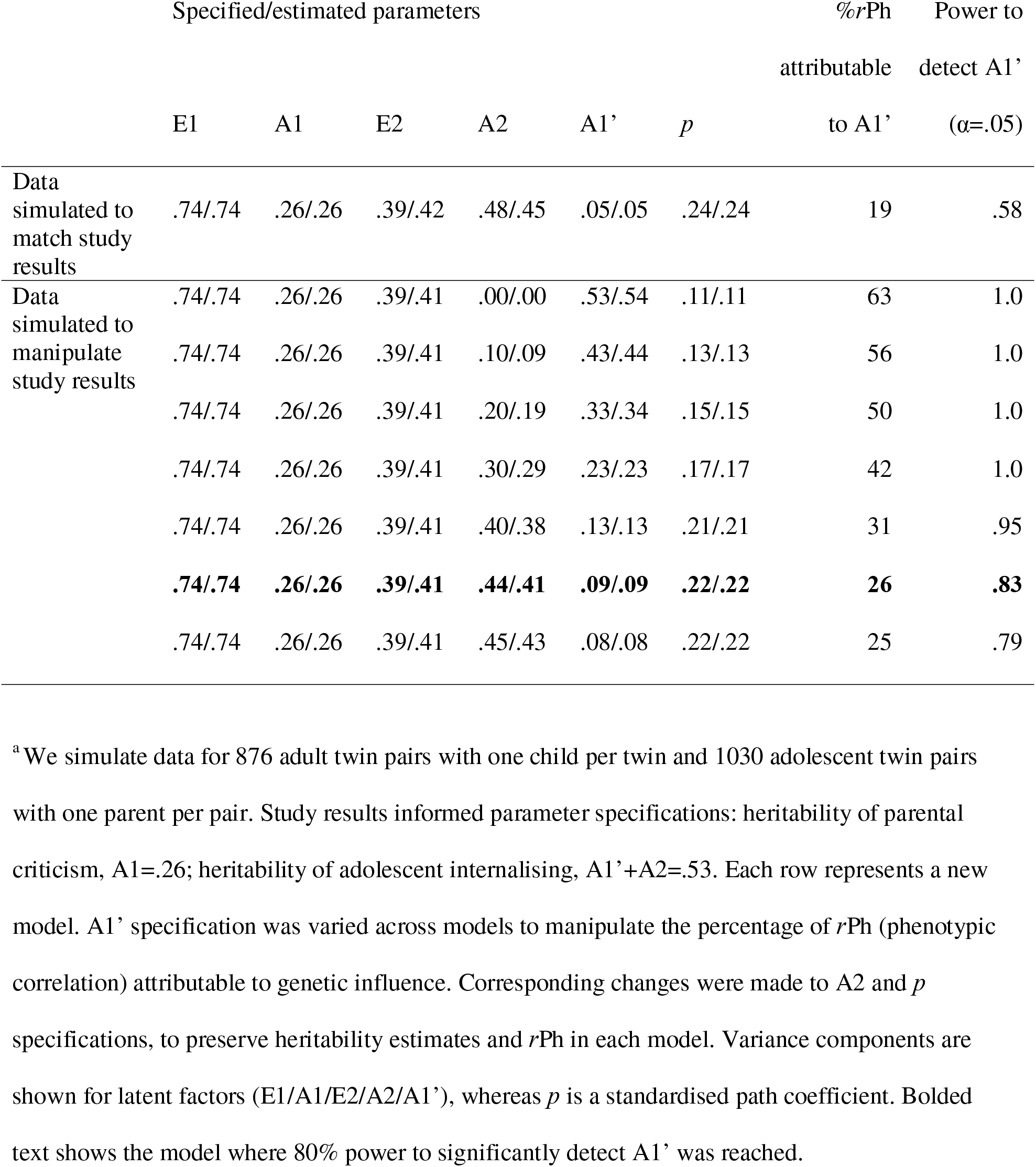
Exploring statistical power to detect genetic influence on the association between parental criticism and adolescent internalising symptoms (*r*=.29)^a^

Statistical power was lower in models using only self-report data for adolescent symptoms, because of the weaker phenotypic parent-child correlation. Here, there was ≥80% power to detect an A1’ estimate accounting for ≥41% of the correlation (TableS4). Therefore, results suggest that *at least* 59% of the association between parental criticism and adolescent-reported internalising symptoms was not attributable to genetic influence.

### Exploring the direction of causation

Given evidence that the parent-offspring correlation was largely attributable to non-genetic influence, we applied causal versions of our Children-of-Twins model, to explore whether non-genetic effects were best modelled as parent-to-offspring and/or offspring-to-parent processes (Heath et al., 1993; Narusyte et al., 2008). In this instance we were unable to distinguish a direction of causation. Details and discussion of our model fitting process and results are included in the supplementary materials. In brief, establishing causality was dependent on our variables having distinct aetiological compositions, which unfortunately was not the case.

## Discussion

Parent reported criticism towards adolescent offspring remained significantly correlated with adolescent internalising symptoms after we accounted for their genetic relatedness. Both parental criticism and offspring internalising symptoms were heritable, but no evidence for genetic covariance between them was found. Specifically, at least 74% of the parent-offspring correlation was attributable to non-genetic effects, which could arise from mechanisms in the immediate family environment. Sensitivity analyses replicated evidence for significant, non-genetic effects, when examining a weaker parent-offspring correlation derived solely from self-reported data (i.e., excluding parent reports of adolescent symptoms). However, genetic effects were more likely to go undetected here, owing to lower statistical power. Further work, drawing on additional data, is needed to explore the causal pathways underpinning non-genetic effects identified between generations.

Previous Children-of-Twins studies examining offspring internalising outcomes conclude that genetic relatedness cannot explain associations with parents’ harsh punishment (Lynch et al., 2006), parent-offspring relationship quality (Hannigan et al., 2018), nor parental criticism (Horwitz et al., 2015). This is surprising, given the heritability of these behaviours and diffuse effects of implicated genes (Plomin et al., 2016). However, previous studies did not include power analyses for the detection of genetic influence, so their findings remain ambiguous. Here we demonstrate that our primary finding (i.e., that the association between parental criticism and adolescent internalising symptoms remains significant after controlling for genetic relatedness) is not attributable to a lack of statistical power. Because previous studies used either the same sample as us (Hannigan et al., 2018; Horwitz et al., 2015), or samples of a similar size (Lynch et al., 2006), our power analyses indicate that their results are likely valid (though this is contingent on comparable phenotypic correlations and heritability estimates). We suggest that the presentation of power analyses in future Children-of-Twins research will aid interpretation of findings (see McAdams et al., 2018, for methodology). Alongside statistical power, we consider how methodological decisions relating to measurement protocol can impact the interpretation of results.

We incorporated data from both parent and offspring reports of adolescent internalising symptoms, which were moderately correlated. Their partial agreement could reflect both shared perspectives and unique insights, alongside perceptual and reporting biases. Adolescents may report symptoms that are unknown to their parents, while parents may identify symptoms that adolescents have not yet recognised (De Los Reyes & Kazdin, 2005). However, using parent reports for all study measures can inflate parent-child correlations via shared method variance. Supplementary analyses using only adolescents’ self-reports of internalising showed consistent findings with our primary analyses, although the phenotypic parent-child correlation was attenuated (.29 to .18). Previously reported correlations are similar to ours, for example *r*=.22-.30 between coded speech samples for parental criticism and maternal reports for adolescent internalising (Frye & Garber, 2005), with lower correlations (*r*=.02-.24) reported when adolescents rate their own symptoms (Nelemans et al., 2014). We show that statistical power to identify genetic confounding is lower for smaller parent-child correlations. When *r*=.18, we had 80% power to detect genetic influence accounting for 59% of the parent-child correlation. When *r*=.29, we had 80% power to detect genetic influence accounting for 74%. Variation in research methodology can ultimately exert influence on the interpretation of results.

Overall, our findings support existing research that has not been genetically informed, suggesting that parental criticism may be relevant to the environment in which adolescent internalising symptoms present (Asarnow et al., 2001; Frye & Garber, 2005; Nelemans et al., 2014; Silk et al., 2009). We note that our results, indicating no genetic correlation between parent and offspring variables, could be in accordance with a finding of genetic nurture in family-based genomic analyses (i.e., environmental parent-to-child effects that occur when parent behaviour is under genetic influence unique to the parent). However, the causal direction of effects could not be determined in our analyses (Supplementary Text2), so remains unknown.

Three complementary study designs could help to identify parent-to-offspring and offspring-to-parent effects. First, child twin studies inform on the extent to which *child* genes influence, or evoke, the parenting they receive (Klahr & Burt, 2014). We found some evidence for adolescent-to-parent processes. Parents’ reports of criticism were almost twice as correlated for MZ (.73) versus DZ (.43) adolescent twins, suggesting that adolescents’ genetically influenced behaviours evoked parental criticism. Further, adult twin data shows that parent-to-child effects are possible, as parents’ genes also influence their parenting behaviours (Klahr & Burt, 2014). To date, the only Children-of-Twins study to directly test the direction of causation between parenting and adolescent internalising found evidence for offspring-to-parent effects (Narusyte et al., 2008), although we present corrections to their model specification and emphasise that their results could be biased by inadequate specification of measurement error (Supplementary Text2). Overall, twin findings do support the possibility of transactional associations between adolescent internalising symptoms and parental criticism.

Second, longitudinal studies have generally identified transactional associations between parental criticism and adolescent internalising symptoms (Frye & Garber, 2005; Nelemans et al., 2014; Silk et al., 2009). One study with six time-frames showed that adolescent internalising was more predictive of parent-reported criticism (using similar questionnaire items as in this study), compared to the reverse (Nelemans et al., 2014). Whilst the association between aspects of the home environment and offspring depression remains relatively stable from childhood into adolescence, the role of offspring genetic influences increases in adolescence. So, adolescents may influence their environments, and thus the parenting they receive, to a greater degree than is seen in younger children (Hannigan, McAdams, & Eley, 2017).

Third, studies that take an experimental or intervention-based approach can help delineate parent-to-child and child-to-parent effects. For example, parental displays of controlling behaviours in experimental settings leave children feeling less capable on a given task, with this effect moderated by child trait anxiety (Thirlwall & Creswell, 2010). Others show that parenting skills training can reduce internalising symptoms among young offspring (Cartwright-Hatton, McNally, White, & Verduyn, 2005), suggesting parent-to-child effects. Conversely, improvements in parenting are observed following treatment of adolescent anxiety (Silverman, Kurtines, Jaccard, & Pina, 2009), suggesting adolescent-to-parent effects. It is important to recognise that the direction of causation in associations between parenting and adolescent outcomes is unlikely to run solely from parents to offspring.

Some study limitations require consideration. Despite our sophisticated research design and large sample, concerns about statistical power meant that we did not examine moderation by participant age and sex. It would be preferable to examine longitudinal data and include two parents per child, to provide a developmental perspective and explore family interactions. Results should be interpreted in the context of our measures, which reflect participants’ own perceptions and are subject to reporter biases, including social desirability bias. Speech samples are sometimes labelled the ‘gold-standard’ for assessing parent expressed criticism (e.g., Asarnow et al., 2001), however these data are rarely available with the sample sizes required for genetically-informed research. Whilst researcher-coded assessments could reduce reporter bias, their results are specific to a window of observation, which may not generalise to parents’ everyday experiences and perceptions. Nevertheless, we had limited data on reliability for our parent measure and note that low reliability may have reduced intergenerational associations. Parent-offspring associations were reduced when using only adolescent self-reports of internalising, which could reflect bias by shared-method-variance in our main results. Future analyses could incorporate data from both reporters separately in the model, to explore variance explained by rater-specific versus common perspectives.

## Conclusion

Parental criticism is associated with the development of adolescent internalising problems. We contextualise previous research, by showing that this association remains significant after we account for genetic influence. We bolster these results with power analyses for the detection of genetic influence. Although we could not distinguish a causal direction for non-genetic effects between generations, literature from child twin, longitudinal and experimental studies all indicate bidirectionality. In sum, parental criticism can be considered part of the environment relevant to the presentation of adolescent internalising symptoms, with adolescent symptoms potentially leading to an increase in parental criticism, and vice versa.

**Key Points**

1. Parental criticism is associated with adolescent internalising problems. The Children-of-Twins design can disentangle genetic and non-genetic influence on this association.
2. Previous Children-of-Twins research suggests that non-genetic influence explains all of the association between parenting behaviours and adolescent internalising. But these studies may have been underpowered to detect genetic influence.
3. Here, the association between parental criticism and adolescent internalising symptoms was attributable to non-genetic influence. Statistical power was sufficient to show that genetic influence accounted for less than 26% of the correlation.
4. There may be causal pathways underlying the correlation between parental criticism and adolescent internalising symptoms. More research is needed to unravel the direction of effects.

## Data Availability

Study data are not publicly available due to privacy/ethical restrictions. Data access is available to researchers on request, subject to data sharing agreements.

## Acknowledgements

This work was supported by a Leverhulme Trust grant awarded to TE (RPG-210). TE is part-funded by a program grant from the UK MRC (MR/V012878/1); NIHR Biomedical Research Centre at South London and Maudsley NHS Foundation Trust; and King’s College London. TM and YA were supported by a Sir Henry Dale Fellowship jointly funded by the Wellcome Trust and the Royal Society (107706/Z/15/Z) and a Wellcome Trust Senior Research Fellowship (220382/Z/20/Z), both awarded to TM. The TOSS was supported by grant R01MH54610 from NIMH (Cohort 1 PI: DR ; Cohort 2 PI: JN). The TCHAD was funded by the Swedish Council for Working Life and Social Research (2004-0383) and Swedish Research Council (2004-1415).

## Footnotes

1 Single-generation, univariate twin model results (available on request) confirmed that variance in adolescent internalising symptoms was not explained by environmental factors shared between twins (C; β=√<.001, 95% CI .00-.12). Therefore, any effects of parental criticism that were shared between twins and not attributable to genetic similarities (alongside other shared environmental influences) were not large enough to be detected as a significant source of variation for adolescent internalising.

2 Calculated by multiplying the paths involved in the genetic route connecting parent criticism to adolescent internalising: a1 (√.26), A1 to A1□ (.5), and a1□ (√.05), divided by the total phenotypic correlation (*r*=.29).

