## Supplementary materials for "Parental criticism and adolescent internalising symptoms: Using a Children-of-Twins design with power calculations to account for genetic influence"

- **Figure S1.** Model specification for decomposing the association between parental criticism and adolescent internalising symptoms

- **Supplementary Text 1.** Notes on model specification for combining adult and child twin datasets

- **Table S1**. Descriptive statistics

- **Table S2.** Correlations in monozygotic (MZ) and dizygotic (DZ) families, using self-report for adolescent internalising symptoms

- **Table S3**. Model fit statistics

- **Figure S2.** Model results, showing the association between parental criticism and offspring (self-reported) internalising symptoms

- **Table S4.** Exploring statistical power to detect genetic influence on the association between parental criticism and adolescent (self-reported) internalising symptoms ($r$=.18)

- **Supplementary Text 2.** Direction of causation for non-genetic, intergenerational effects

- **Figure S3.** Model specification for a two-sample direction of causation Children-of-Twins model, used to decompose the association between parental criticism and adolescent internalising symptoms

- **Table S5.** Standardised parameter estimates (95% confidence intervals) and model fit in two-sample direction of causation Children-of-Twins models

**Figure S1.** Model specification for decomposing the association between parental criticism and adolescent internalising symptoms [a]

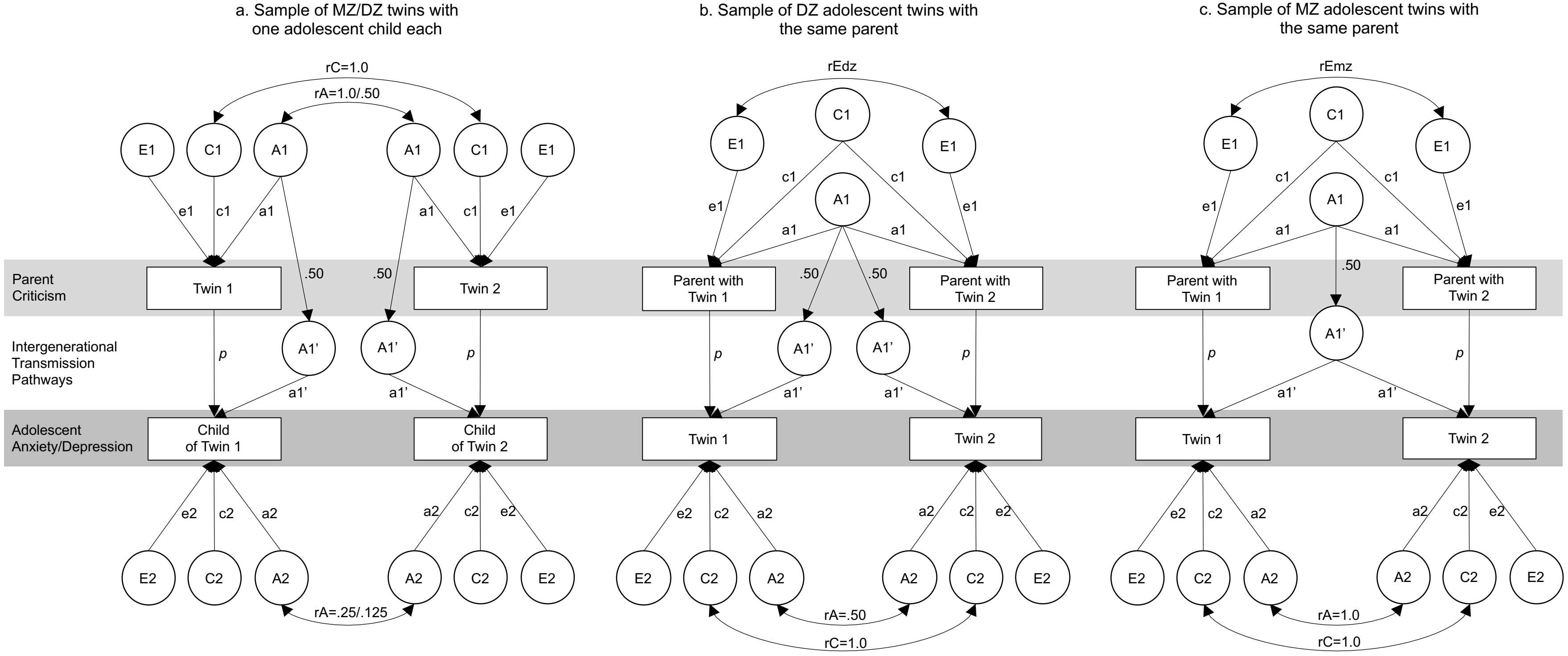


[a] A1=additive genetic effects on parent criticism; C1=shared-environmental effects on parent criticism; E1=nonshared environmental effects on parent criticism; A2= additive genetic effects specific to adolescent internalising symptoms; C2=shared-environmental effects on adolescent internalising symptoms (cannot be estimated using cousin data); E2=nonshared environmental effects on adolescent internalising symptoms; A1′=genetic effects common to parent criticism and adolescent internalising symptoms; p=residual non-genetic association between parental criticism and adolescent internalising symptoms; rEmz/rEdz=freely estimated correlations to allow the parenting of adolescent Twin 1 and Twin 2 to differ from one another, while also ensuring that these within-parent correlations can differ across adolescent zygosity (panels b and c) and can differ from adult MZ twin correlations (panel a). The pathway between A1 and A1′ is fixed to .50 because children inherit 50% of their parent's genes. Variance=1 for all latent factors.

**Supplementary Text 1.** Notes on model specification for combining adult and child twin datasets

We specified data from DZ and MZ adolescent twins separately, to reflect that MZ twins inherit the same genes from their parent (FigureS1, panel 'c'), whereas DZ twins each inherit a random 50% (FigureS1, panel 'b'). This means that covariance between adolescent MZ twins includes A1', whereas covariance between DZ twins includes .25*A1'. This specification was not included in previously published studies (e.g., [1]).

Further, we included parameters rEdz and rEmz to distinguish 'within-parent' correlations in the adolescent twin sample from MZ parent correlations in the adult twin sample.[2] This distinguishes individual parents in the adolescent twin sample from genetically identical parent pairs in the adult twin sample. rEdz and rEmz also allow the 'within-parent' correlations to differ between parents of MZ and DZ adolescent twins, to account for differences in evocative effects from MZ and DZ pairs (evocative child effects can make the parenting of MZ twins more correlated than the parenting of DZ twins). In some previous studies, correlations between parents who are MZ twin pairs (FigureS1, top of panel 'a'); within-parent correlations for parenting of DZ twins (FigureS1, top of panel 'b'); and within-parent correlations for parenting of MZ twins (FigureS1, top of panel 'c') were all constrained to be the same (A1+C1). In our model specification they can all vary. This means that in our model, A1 and C1 estimates are derived from the Children-of-Twins sample only. Within-parent correlations in the adolescent twin sample are not brought into the estimation of parent aetiology.

1. Narusyte J, Neiderhiser JM, D'Onofrio BM, et al. Testing different types of genotype-environment correlation: an extended children-of-twins model. *Dev Psychol.* 2008;44(6):1591-1603.
2. McAdams TA, Hannigan LJ, Eilertsen EM, Gjerde LC, Ystrom E, Rijsdijk FV. Revisiting the Children-of-Twins Design: Improving Existing Models for the Exploration of Intergenerational Associations. Behavior Genetics. 2018;48(5):397-412.

**Table S1**. Descriptive statistics
[a] Expressed Emotion measure. [b] Child Behaviour Checklist. [c] Youth Self-Report.

| Sample | Reporter (N items) | Cronbach's alpha | N | Raw data / Log transformed data | | | |
|---|---|---|---|---|---|---|---|
| | | | | Mean (SD) | Variance | Skew | Kurtosis |
| *Parental criticism* | | | | | | | |
| Children of Twins (TOSS) | Parent (10) [a] | .86 | 1,721 | 17.4 (5.26)/2.16 (0.54) | 27.7/0.30 | 0.98/-0.33 | 3.83/2.85 |
| Adolescent Twins (TCHAD) | Parent (10) [a] | .90 | 2,112 | 16.9 (5.89)/0.65 (0.27) | 34.7/0.07 | 1.07/0.51 | 3.92/2.50 |
| *Offspring internalising symptoms* | | | | | | | |
| Children of Twins (TOSS) | Parent (30) [b] | .81 | 1,706 | 3.83 (4.21)/1.65 (0.61) | 17.8/0.37 | 2.17/0.28 | 10.5/2.70 |
| | Child (30) [c] | .86 | 1,669 | 8.70 (6.62)/2.35 (0.55) | 43.8/0.30 | 1.35/-0.57 | 5.30/4.44 |
| | Composite | -- | 1,743 | 6.26 (4.63)/1.81 (0.52) | 21.4/0.27 | 1.50/-0.20 | 5.87/3.35 |
| Adolescent Twins (TCHAD) | Parent (30) [b] | .80 | 2,087 | 3.40 (4.50)/1.60 (0.60) | 20.3/0.36 | 2.55/0.40 | 11.3/3.14 |
| | Child (30) [c] | .88 | 2,313 | 8.34 (7.14)/2.33 (0.58) | 50.9/0.34 | 1.35/-0.49 | 5.02/3.69 |
| | Composite | -- | 2,420 | 6.16 (5.48)/2.07 (0.51) | 30.0/0.26 | 1.77/-0.19 | 7.46/3.39 |

**Table S2.** Correlations in monozygotic (MZ) and dizygotic (DZ) families, using self-report for adolescent internalising symptoms

| | MZ twin families | DZ twin families |
|---|---|---|
| *Parental criticism* | | |
| Twin parent correlations on parental criticism for adolescent offspring [a] | .25 (.14, .34) | .14 (.09, .22) |
| *Parental criticism and self-report offspring internalising symptoms* | | |
| Parent-child correlation [b] | .18 (.15, .22) | .18 (.15, .22) |
| Avuncular correlations [a] | .07 (.02, .12) | .04 (.01, .06) |
| *Self-report offspring internalising symptoms* | | |
| Cousin correlations on internalising symptoms [a] | .13 (.09, .16) | .07 (.05, .08) |
| Twin correlations on internalising symptoms [c] | .50 (.42, .58) | .26 (.19, .32) |

[a] Correlations taken from the Children of Twins sample, TOSS. [b] Correlations taken from TOSS *and* the adolescent twin sample, TCHAD. [c] Correlations taken from TCHAD. Brackets show 95% confidence intervals.

**Table S3**. Model fit statistics

| | | -2LL | df | AIC | Compare to full model Δ-2LL | Δ df | *p* | Compare to AE model Δ-2LL | Δ df | *p* |
|---|---|---|---|---|---|---|---|---|---|---|
| Models using composite score for adolescent internalising symptoms | Full model (ACE model) | 20361 | 7545 | 5271 | - | - | - | - | - | - |
| | No C paths (AE model) | 20361 | 7547 | 5267 | .08 | 2 | .96 | - | - | - |
| | AE model with no A1’ path | 20365 | 7548 | 5269 | 3.9 | 3 | .28 | 3.8 | 1 | .05 |
| | AE model with no p path | 20440 | 7548 | 5344 | 79 | 3 | <.001 | 79 | 1 | <.001 |
| Models using self-report for adolescent internalising symptoms | Full model (ACE model) | 20187 | 7368 | 5451 | - | - | - | - | - | - |
| | No C paths (AE model) | 20187 | 7370 | 5447 | .11 | 2 | .95 | - | - | - |
| | AE model with no A1’ path | 20188 | 7371 | 5446 | .80 | 3 | .85 | .69 | 1 | .41 |
| | AE model with no p path | 20212 | 7371 | 5470 | 24 | 3 | <.001 | 24 | 1 | <.001 |

**Figure S2.** Model results, showing the association between parental criticism and offspring (self-reported) internalising symptoms [a]

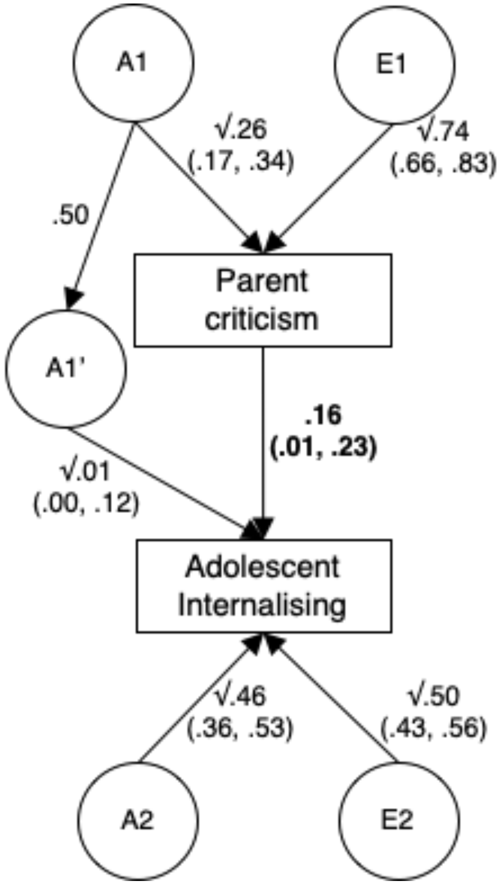


[a] A1=additive genetic effects on parent trait; A2=additive genetic effects specific to adolescent trait; A1′=additive genetic effects common to parent and adolescent traits (path from A1 to A1′ is fixed to .50 because offspring inherit 50% of their parent's genes); C1/C2=shared environment effects on parent/adolescent trait; E1/E2=nonshared environmental effects on parent/adolescent trait. Variance components are displayed, with the residual intergenerational association in bold (standardised path beta coefficient). The relative contribution of A1' in explaining the phenotypic parent-child correlation is calculated by multiplying the genetic paths connecting each trait, divided by the total phenotypic correlation ($r$=.18).

**Table S4.** Exploring statistical power to detect genetic influence on the association between parental criticism and adolescent (self-reported) internalising symptoms (*r*=.18) [a]

| | Specified/estimated parameters | | | | | | %*r*Ph attributable to A1' | Power to detect A1' (α = .05) |
|---|---|---|---|---|---|---|---|---|
| | E1 | A1 | E2 | A2 | A1' | *p* | | |
| Data simulated to match study results | .74/.74 | .26/.26 | .50/.51 | .46/.45 | .01/.01 | .16/.16 | 14 | .15 |
| Data simulated to manipulate study results | .74/.74 | .26/.26 | .50/.51 | .00/.00 | .47/.48 | .01/.01 | 95 | 1.0 |
| | .74/.74 | .26/.26 | .50/.51 | .10/.11 | .37/.38 | .03/.03 | 84 | 1.0 |
| | .74/.74 | .26/.26 | .50/.51 | .20/.20 | .27/.27 | .05/.05 | 73 | 1.0 |
| | .74/.74 | .26/.26 | .50/.51 | .30/.30 | .17/.17 | .08/.08 | 57 | .98 |
| | .74/.74 | .26/.26 | .50/.51 | .35/.35 | .12/.12 | .10/.10 | 47 | .90 |
| | .74/.74 | .26/.26 | .50/.51 | .37/.37 | .10/.10 | .10/.10 | 45 | .84 |
| | **.74/.74** | **.26/.26** | **.50/.51** | **.38/.37** | **.09/.09** | **.11/.11** | **41** | **.80** |
| | .74/.74 | .26/.26 | .50/.51 | .40/.40 | .07/.07 | .11/.11 | 38 | .69 |

[a] We simulate data for 876 adult twin pairs with one child per twin and 1030 adolescent twin pairs with one parent per pair. Parameters are specified based on study results, where parental criticism heritability was .26 (A1) and adolescent internalising heritability was .47 (A1' + A2). Each row represents a new model. A1' specification is varied across models to manipulate the percentage of rPh (phenotypic correlation) attributable to genetic confounding. Corresponding changes are made to A2 and *p* specification, to preserve heritability estimates and rPh in each model. Latent factors have variance components (E1/A1/E2/A2/A1'), whereas p is a standardised path coefficient. Bolded text shows the model where 80% power to significantly detect A1' was reached.

**Supplementary Text 2.** Direction of causation for non-genetic, intergenerational effects

As a follow-up analysis to the presented Children-of-Twins model, we ran a direction of causation model that included causal paths running from offspring-to-parent and parent-to-offspring (Figure S3). The logic underlying direction of causation models has been discussed elsewhere,[1] as has the application of these models to combined Children-of-Twins and child twin data.[2]

In short, direction of causation can be tested using cross-sectional family data for variables that differ in their aetiology (i.e., relative magnitudes of direct genetic and shared environmental influences).[1,3] When there is a causal relationship between two traits, and these traits differ in their aetiology, the covariance between the traits can be used to determine the direction of causation. For example, if X causes Y, and X is more heritable than Y, then the covariance between X and Y will be driven by genetic factors. As such, the covariance decomposition between X and Y can be used to trace the origins of their association. Put another way, the variance components of X can be thought of as instrumental variables in their prediction of Y, as they only become associated with Y via causal influence of X on Y.[4]

We adapted the model specification introduced by Narusyte et al., where causal intergenerational paths run from parent-to-offspring (m) *and* offspring-to-parent (n) traits, alongside the A1' path for genetic transmission (Figure S3).[2] We made the same amendments to the model specification as discussed in Supplementary Text 1 for the standard model (i.e., we include rEmz and rEdz parameters; and specify the model to ensure that MZ twin children share an identical A1' factor). Further, we ran several iterations of the model to account for possible bias introduced by measurement error.

Measurement error is an important consideration in all tests of causality between variables. If significant measurement error is present (as is the norm in behavioural science, where variables are rarely measured without considerable error), then this must be modelled to avoid biasing other parameter estimates.[1,3] In direction of causation twin models, measurement error ($\varepsilon$) is not subsumable as unshared environmental variation (E), as is the case in the standard factor models.[3] When researchers lack the data required to specify a value for $\varepsilon$ a priori (i.e., when measurement error is unknown, as was the case in our analyses), then $\varepsilon$ should be freely estimated within the model. In direction of causation Children-of-Twins models, it is not possible to estimate $\varepsilon$ separately for two variables alongside the causal paths m and n – this model (as shown in Figure S3) is not identified.

Narusyte et al. incorporate a freely estimated $\varepsilon$ parameter in their reciprocal direction of causation Children-of-Twins model, with $\varepsilon$ contributing directly to variance in both generations.[2] Their model is identified because $\varepsilon$ is fixed to be equal across both variables (i.e., fixing $\varepsilon 1$ and $\varepsilon 2$ to equality in Figure S3). However, the validity of this method is undermined if true measurement error differs between variables. To avoid this pitfall, we followed a protocol suggested by Heath et al., who showed that measurement error need only be estimated for the *predictor* variable in unidirectional causal models (i.e., estimating only $\varepsilon 1$ if modelling only a parent-to-offspring causal path; and estimating only $\varepsilon 2$ if modelling only an offspring-to-parent path), and doing so gives an unbiased estimate of the causal influence of predictor on outcome.[1] We ran sequential univariate causal models to derive parameter estimates that were not biased. Results are presented in Table S5, panel A, and show that our data were explained equally well by causal paths in either direction. Model fit statistics provided no clear reason to choose one model over the other. Because these models showed overlapping confidence intervals for $\varepsilon$ estimates in both generations, we next followed the Narusyte et al. method of fixing $\varepsilon$ estimates in both generations to equality, to simultaneously estimate parent-to-child and child-to-parent effects. Results are provided in Table S5, panel B, and show that neither causal path was estimated as significantly different from zero. Further, it was possible to drop either the parent-to-offspring (m) *or* the offspring-to-parent (n) paths from this model without compromising model fit.

By applying these tests regarding the direction of causality, we hoped to identify whether the non-genetic association between parental criticism and offspring internalising symptoms could best be conceptualised as a unidirectional parent-to-offspring or offspring-to-parent effect, or by bidirectional effects in both directions. We found no clear evidence to help in differentiating between these possibilities (Table S5).

There are a number of interrelated possibilities that may explain our inability to distinguish direction of causation. Primarily, the aetiological structures of our phenotypes were likely too similar to distinguish reciprocal causation.[1,3] Although adolescent internalising symptoms were more heritable than parental criticism, their overall aetiologies were perhaps not different enough to be distinguishable in our sample. This is shown by the overlapping confidence intervals for parent and offspring trait heritability (A1 and A2) in the bidirectional model (Table S5, panel B). It may have been easier to distinguish the 'm' and 'n' paths if C2 were significant, because C2 would impact upon the parent-offspring covariance structure via the offspring-to-parent 'n' path. Further, variance explained by measurement error ($\varepsilon$) did not differ significantly between variables, again adding to their homogenous composition. As demonstrated via previously published power calculations, statistical power to establish causality depends on the relative degrees of measurement error in both variables, as well as the underlying aetiological structure of the phenotype.[1,2] Finally, causality between variables, in either direction, is a major assumption in direction of causation analyses.[4] Although we show that the association between parental criticism and adolescent internalising symptoms was not attributable to genetic factors in our sample, influence by common causes in the nuclear family environment (e.g., neighbourhood stressors, socio-economic factors, or other family members) remained possible. Influence by non-genetic confounders may have contributed to our inability to distinguish intergenerational causal paths.

1. Heath AC, Kessler RC, Neale MC, Hewitt JK, Eaves LJ, Kendler KS. Testing hypotheses about direction of causation using cross-sectional family data. *Behav Genet.* 1993;23(1):29-50.
2. Narusyte J, Neiderhiser JM, D'Onofrio BM, et al. Testing different types of genotype-environment correlation: an extended children-of-twins model. *Dev Psychol.* 2008;44(6):1591-1603.
3. Duffy DL, Martin NG. Inferring the direction of causation in cross-sectional twin data: theoretical and empirical considerations. *Genet Epidemiol.* 1994;11(6):483-502.
4. McAdams TA, Rijsdijk FV, Zavos HM, Pingault J-B. Twins and causal inference: Leveraging nature's experiment. *Cold Spring Harbor Perspectives in Medicine.* 2020.

**Figure S3.** Model specification for a two-sample direction of causation Children-of-Twins model, used to decompose the association between parental criticism and adolescent internalising symptoms [a]

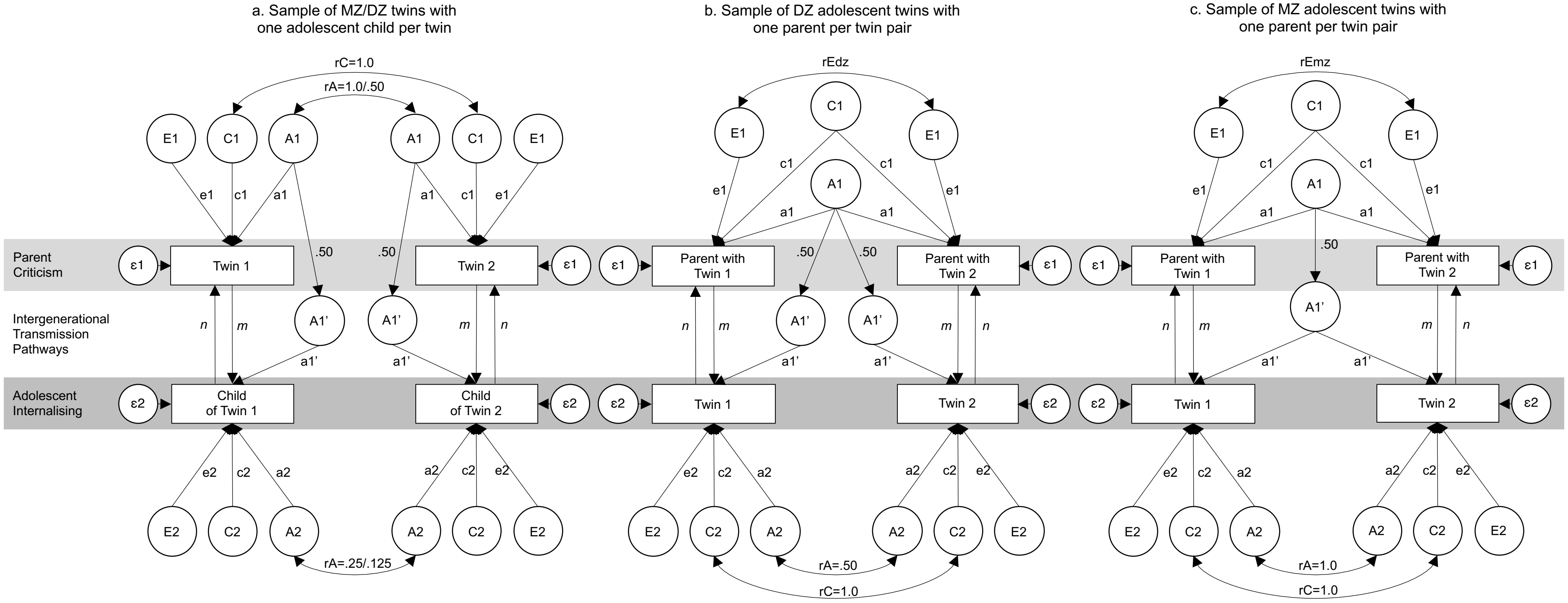


[a] A1=Additive genetic effects on parental phenotype; C1= shared-environmental effects on parental phenotype; E1=nonshared environmental effects on parental phenotype; A1'=genetic effects common to parental phenotype and offspring phenotype; A2=genetic effects specific to offspring phenotype; C2= shared-environmental effects on offspring phenotype (not estimable using cousin data); E2=nonshared environmental effects on offspring phenotype; m=phenotypic effect of parent on offspring; n=phenotypic effect of offspring on parent; rEmz/rEdz=freely estimated correlations to allow the parenting of adolescent Twin 1 and Twin 2 to differ from one another, while also ensuring that these within-parent correlations can differ across adolescent zygosity (panels b and c) and can differ from adult MZ twin correlations (panel a). Measurement error (ε1 and ε2) contributes directly to phenotype variance. **Note that this model is not identified with all parameters freely estimated.** See Supplementary Text 2 for further discussion.

**Table S5.** Standardised parameter estimates (95% confidence intervals) and model fit in two-sample direction of causation Children-of-Twins models [a]

**A. Testing unidirectional causation: measurement error estimated for the predictor variable**

| | A1 | C1 | E1 | A2 | C2 | E2 | A1’ | ε1 | ε2 | m | n | -2LL (df) | AIC |
|---|---|---|---|---|---|---|---|---|---|---|---|---|---|
| Child-to-parent | .36 (.27, .44) | - | .64 (.56, .73) | .61 (.56, .66) | - | .03 (.00, .10) | - | - | .36 (.27, .43) | - | .46 (.39, .54) | 20355.10 (7547) | 5261.10 |
| Parent-to-child | .27 (.18, .35) | - | .52 (.41, 64) | .55 (.49, .60) | - | .45 (.40, .51) | - | .21 (.12, .29) | - | .38 (.33, .44) | - | 20353.05 (7547) | 5259.05 |

**B. Testing bidirectional causation: measurement error estimated as equal for both variables**

| | A1 | C1 | E1 | A2 | C2 | E2 | A1’ | ε | m | n | -2LL (df) | AIC | Δ-2LL (Δdf) | p |
|---|---|---|---|---|---|---|---|---|---|---|---|---|---|---|
| Bidirectional | .27 (.18, .43) | - | .51 (.16, .79) | .55 (.40, .65) | - | .23 (.00, .60) | - | .22 (.00, .42) | .37 (-.15, .52) | .01 (-.38, .56) | 20353.05 (7546) | 5261.05 | | |
| Child-to-parent | .36 (.27, .44) | - | .28 (.15, .41) | .61 (.56, .66) | - | .03 (00, .10) | - | .36 (.28, .44) | - | .46 (.39, .54) | 20355.10 (7547) | 5261.10 | 2.05 (1) | .15 |
| Parent-to-child | .27 (.18, .35) | - | .52 (.41, .60) | .55 (.49, .60) | - | .24 (.16, .34) | - | .21 (.12, .29) | .38 (.33, .44) | - | 20353.05 (7547) | 5259.05 | 0.00 (1) | .97 |

[a] A1=Additive genetic effects on parental criticism; C1= shared-environmental effects on parental criticism; E1=nonshared environmental effects on parental criticism; A2=genetic effects specific to offspring internalising; C2= shared-environmental effects on offspring internalising (not estimable using cousin data); E2=nonshared environmental effects on offspring internalising; A1’=genetic effects common to parental phenotype and offspring internalising; ε=measurement error; m=non-geneitc effect of parent on offspring; n=non-genetic effect of child on parent. Parameters C1, C2 and A1’ did not explain signficant variance and were subsequently fixed to zero, as denoted by empty cells (-), without significant change to model fit. In Panel A, child-to-parent causation is tested by estimating ε independently in the child generation (ε2); then parent-to-child causation tested by estimating ε independently in the parent generation (ε1). In Panel B, model identification is achieved for testing bidirectional causation by fixing ε to unity across generations. This is justified by results in panel A, showing overlapping confidence intervals for ε1 and ε2. All results suggest that data are explained equally well by child-to-parent or parent-to-child causation. We find no clear evidence for differentiating the two. See Supplementary Text 2 for further discussion.
